# Long COVID and financial outcomes: Evidence from four longitudinal population surveys

**DOI:** 10.1101/2023.05.23.23290354

**Authors:** Rebecca Rhead, Jacques Wels, Bettina Moltrecht, Richard J. Shaw, Richard J. Silverwood, Jingmin Zhu, Alun Hughes, Nishi Chaturvedi, Evangelia Demou, Srinivasa Vittal Katikireddi, George B. Ploubidis

## Abstract

**Background:** Long-term sequelae of COVID-19 (long COVID) include muscle weakness, fatigue, breathing difficulties and sleep disturbance over weeks or months. Using UK longitudinal data, we assessed the relationship between long COVID and financial disruption.

**Methods:** We estimated associations between long COVID (derived using self-reported length of COVID-19 symptoms) and measures of financial disruption (subjective financial well-being, new benefit claims, changes in household income) by analysing data from four longitudinal population studies, gathered during the first year of the pandemic. We employed modified Poisson regression in a pooled analysis of the four cohorts adjusting for a range of potential confounders, including pre-pandemic (pre-long COVID) factors.

**Results:** Among 20,112 observations across four population surveys, 13% reported having COVID-19 with symptoms that impeded their ability to function normally - 10.7% had such symptoms for <4 weeks (acute COVID-19), 1.2% had such symptoms for 4-12 weeks (ongoing symptomatic COVID-19) and 0.6% had such symptoms for >12 weeks (post-COVID-19 syndrome). We found that post-COVID-19 syndrome was associated with worse subjective financial well-being (adjusted relative risk ratios (aRRR)=1.57, 95% confidence interval (CI)=1.25, 1.96) and new benefit claims (aRRR=1.79, CI=1.27, 2.53). Associations were broadly similar across sexes and education levels. These results were not meaningfully altered when scaled to represent the population by age.

**Conclusions:** Long COVID was associated with financial disruption in the UK. If our findings reflect causal effects, extending employment protection and financial support to people with long COVID may be warranted.

## Introduction

Many studies in hospitalised and community settings have identified that those with COVID-19 can continue to experience long-term symptoms such as muscle weakness, fatigue, breathing difficulties and sleep disturbance (1). Pooled global prevalence of post-COVID-19 conditions is estimated to be 0.43%(3) while recent estimates from the United States suggest a higher prevalence of 7.5% (2). Such extended COVID-19 symptomatology over weeks or months has been termed ‘long COVID’.

The UK’s National Institute for Health and Care Excellence divides the duration of COVID-19 symptoms into three categories: less than 4 weeks (acute COVID-19), 4-12 weeks (ongoing symptomatic COVID-19), and more than 12 weeks (post-COVID syndrome), with long COVID encompassing the latter two categories (4). In the UK, long COVID prevalence estimates range from 13% in highly selected, community-based survey respondents with test-confirmed SARS-CoV-2, to approximately 71% among those hospitalised by the infection (5–8). Given the scale of the pandemic, even a low proportion of individuals with long COVID will generate a major burden of enduring illness (9,10).

Some long COVID symptoms can hinder an individual’s ability to work, such as post-viral chronic fatigue, cardiopulmonary symptoms, anxiety and depression (11,12). As poor health and disability carry penalties for income and employment (13–15), the COVID-19 pandemic represents a potentially serious threat to both health and prosperity. A recent national registry-based study in Sweden of those receiving sickness benefits for COVID-19 between March-August 2020 found that more than one in ten subjects were on sick leave for over 12□weeks (indicating potential post-COVID syndrome). 13% of those receiving sickness benefits due to COVID-19 were on sick leave for long COVID (16). Long COVID might also have a financial impact on those who are not employed as managing symptoms can incur additional costs (medical costs, paying for additional care or support) and potentially delay or prevent re-entering the job market.

Research into how COVID-19 symptoms and chronicity have disrupted employment is ongoing. A number of studies have demonstrated that individuals who contracted mild or even asymptomatic cases are experiencing lasting symptoms with implications for their day-to-day lives, including their ability to work (17,18). An international, patient-led survey found that 45% of individuals with long COVID had reduced their workload and 22% were not working at the time of the survey due to their COVID-19-related health conditions (19). Findings from the Opinions and Lifestyle Survey (20) conducted by the Office of National Statistics in Great Britain between April-June 2021, found that long COVID was associated with a negative impact on household finances (21) - 44% of survey participants with long COVID said their work had been affected by the pandemic and 22% reported their household finances had been affected in some way. In PHOSP-COVID, a multi-centre UK study of adults hospitalised due to COVID-19, only 29% felt fully recovered, 20% had a new disability, and 19% experienced a health-related change in occupation up to seven months post-discharge (22). However, little is known about whether long COVID is associated with increased use of welfare benefits or a reduction in income.

Any financial disruption associated with COVID-19 is unlikely to impact everyone equally as underlying financial and health inequalities along dimensions such as sex and education may exacerbate the financial impact of ongoing COVID-19 symptoms (23–25). Furthermore, adults with pre-existing mental health conditions are at an increased risk of contracting COVID-19 and are more likely to suffer from worse physical and mental health outcomes following infection (26–28). Further research is needed to understand the effects of long COVID on financial wellbeing (29), particularly as many countries are entering a period of recession and cost of living pressures. We use data from four UK longitudinal population studies with rich pre-pandemic longitudinal data to investigate whether experiencing long COVID is associated with financial disruption and if these associations are modified by sex and education.

## Methods

### Data

Data for this study come from four UK cohorts:

- **Millennium Cohort Study (MCS)** (30) - 18,818 people born in the UK in the year 2000–2001.
- **Next Steps (NS, formerly the Longitudinal Study of Young People in England)** (31) - 16,000 people in England born in 1989-90.
- **1970 British Cohort Study (BCS70)** (32) - 17,000 people born in England, Scotland and Wales in a single week of 1970.
- **1958 National Child Development Study (NCDS)** (33) - 17,415 people born in England, Scotland and Wales in a single week of 1958.

Cohort members were asked to participate in a series of COVID-19 surveys to understand the economic, social and health impacts of the COVID-19 crisis. Three waves of this COVID-19 survey were conducted between April 2020 and March 2021 (34).

### Exposure

Our exposure was long COVID which was derived from self-reported data from the 3^rd^ wave of the COVID-19 survey (February – March 2021) on the duration of symptoms following COVID-19. Participants were asked first if they had ever had COVID-19, if they indicated ‘yes, confirmed by a positive test’ or ‘yes, based on strong personal suspicion or medical advice’, they were asked how long they were unable to function as normal due to COVID-19 symptoms. From these responses, the following long-COVID categories were derived, reflecting acute, ongoing symptomatic and post-COVID19 syndrome. This measure allowed us to understand the financial impact of COVID-19 symptoms duration.

0 No SARS-CoV-2/COVID-19 (reference category)
1 SARS-CoV-2 infection with no symptoms / mild symptoms that did not impede normal functioning
2 COVID-19 with symptoms which last less than 4 weeks (acute COVID-19)
3 COVID-19 with symptoms which last longer than 4 weeks but less than 12 weeks (ongoing symptomatic COVID-19)
4 COVID-19 with symptoms which last longer than 12+ weeks (post-COVID-19 syndrome)

Additional sensitivity analysis was conducted to determine how many people reported infection <12 weeks prior to the 3^rd^ wave of the COVID-19 survey and whether their exclusion from analysis impacts results.

### Outcomes

This study has three outcomes, i) subjective financial well-being, ii) new benefits claims during the pandemic, and iii) change in household income, all taken from the 3^rd^ wave of the COVID-19 survey (February – March 2021). Subjective financial well-being was assessed using answers to the question: “Overall, how do you feel your current financial situation compares to before the Coronavirus outbreak in March 2020?”. Responses were measured on a 5-point Likert scale (much worse off – much better off) and dichotomised for this study (worse off = 1, same/better off = 0). Participants were also asked if they had made any new benefit claims since March 2020 (yes/no). These benefits include Universal Credit, free school meals, employment and support allowance, sick pay, council tax support, COVID-19 self-employment support scheme, and career allowance.

Finally, change in income was assessed using data derived from reported weekly household income (after tax) taken at the 3rd wave of the COVID-19 survey (February - March 2021) and pre-pandemic weekly household income (retrospectively assessed at the 3rd wave). Using these data, a binary measure of income change was derived (1 if decreased by at least 5%, and 0 otherwise). Additional sensitivity analysis was also conducted to examine Organisation for Economic Co-operation and Development (OECD) (35) equalised pandemic weekly household income outcome after adjusting for equalised pre-pandemic income.

### Potential confounders

In adjusted models we accounted for sociodemographic characteristics which include sex (male, female), ethnicity (White, non-White (due to low numbers of minority ethnic people within the studies)), keyworker status (yes, no) as well as pre-pandemic employment status (employed, unemployed, economically inactive) and education (degree, no-degree). Adjustments were also made for whether participants shielded during the pandemic (yes, no), if they had any chronic health conditions before the pandemic (yes, no), and pre-pandemic psychological distress (yes, no). Psychological distress was measured using the GHQ-12 (cut off of 4+) in MCS and NS cohorts, and the Malaise Inventory (9-item version, cut off of 4+) for BCS70 and NCDS (36). See Supplementary Material for details of these confounders and a directed acyclic graph to illustrate their relationships with exposure and outcomes.

### Analysis

In this study, we conducted a pooled analysis across multiple cohorts. Descriptive statistics (frequencies and percentages) were reported for all exposures and outcomes by study. Modified Poisson regression with robust standard errors (that return risk ratios) were used to examine subjective financial well-being, new benefit claims and our binary measure of income change. This method was used for ease of interpretation and to avoid issues related to the non-collapsibility of odds ratios (37,38). Due to decreased sample size as a result of missing income data (n=4,760 had missing data for both pre-and post-C19 household income) a four-category measure of long COVID was used when assessing change in income where symptoms lasting between 4-12 weeks (ongoing symptomatic COVID-19) and 12+ weeks (post-COVID-19 syndrome) were combined into a single category (symptoms 4+ weeks).

Crude and adjusted estimates for all models were reported. Missing data for covariates were imputed using multiple imputation with chained equations (39). All potential confounders were included in the imputation models. To examine whether sex and level of education moderated the strength of the associations between long COVID and each outcome, regression models including and excluding interaction terms were fitted and compared using a Wald test (analytic models only).

For the main analyses, cohorts were weighted to account for sampling design and differential non-response. Sampling strata were accounted for and a finite population correction was applied where appropriate (34). In addition, analyses were repeated using weights that scaled each cohort to reflect the actual size they would have in the total population. These population age composition weights were generated using Office for National Statistics data (40) mid-year population estimates by age in 2020 to recalculate the representativeness of each cohort based on the actual distribution in the overall population. All proportions, estimates and 95% confidence intervals (CI) reported in this study were weighted, and frequencies were unweighted. All analyses were conducted using R V.4.2.0. (41).

## Results

Analysis was conducted on a total sample size of N=20,112 (NCDS = 6,467, BCS = 5,421, NS = 4,005, MCS = 4,219). This analytical sample was restricted to non-missing on our long COVID measure as well as subjective financial well-being (n=433 missing) and new benefit claims. As shown in Table 1, 13% of the sample reported having COVID-19 with symptoms that impeded their ability to function normally.

**Table 1:** Exposure and outcomes overall and for each specific cohort.

| Characteristic | Overall<br>N = 20,112<sup>1</sup> | NCDS<br>N = 6,467<sup>1</sup> | BCS70<br>N = 5,421<sup>1</sup> | NS<br>N = 4,005<sup>1</sup> | MCS<br>N = 4,219<sup>1</sup> |
| --- | --- | --- | --- | --- | --- |
| **Long COVID** |  |  |  |  |  |
| No covid | 16,731 (83.1%) | 5,783 (89.4%) | 4,565 (84.2%) | 3,190 (80.6%) | 3,193 (75.3%) |
| C19 - normal functioning | 879 (4.5%) | 133 (2.1%) | 192 (3.5%) | 194 (4.7%) | 360 (8.7%) |
| C19 - sym <4weeks | 2,139 (10.7%) | 426 (6.6%) | 537 (9.9%) | 550 (12.9%) | 626 (15.1%) |
| C19 - sym 4-<12 weeks | 251 (1.2%) | 92 (1.4%) | 82 (1.5%) | 49 (1.1%) | 28 (0.7%) |
| C19 - sym 12+ weeks | 112 (0.6%) | 33 (0.5%) | 45 (0.8%) | 22 (0.6%) | 12 (0.3%) |
| **Coping financially compared to pre-pandemic** |  |  |  |  |  |
| Same/better | 14,585 (72.8%) | 4,740 (73.3%) | 3,940 (72.7%) | 2,932 (73.7%) | 2,973 (71.2%) |
| Worse | 5,527 (27.2%) | 1,727 (26.7%) | 1,481 (27.3%) | 1,073 (26.3%) | 1,246 (28.8%) |
| **New benefit claims since pandemic** |  |  |  |  |  |
| No | 17,303 (86.3%) | 5,637 (87.2%) | 4,631 (85.4%) | 3,356 (84.1%) | 3,679 (88.0%) |
| Yes | 2,809 (13.7%) | 830 (12.8%) | 790 (14.6%) | 649 (15.9%) | 540 (12.0%) |
| **Change in weekly household income since pandemic** |  |  |  |  |  |
| same/increased & decrease < 5% | 12,662 (81.9%) | 3,918 (82.1%) | 3,453 (81.4%) | 2,843 (82.8%) | 2,448 (81.2%) |
| decreased by ≥ 5% | 2,733 (18.1%) | 856 (17.9%) | 787 (18.6%) | 561 (17.2%) | 529 (18.8%) |
| Unknown | 4,760 | 1,693 | 1,181 | 583 | 1,303 |

<sup>1</sup> unweighted n (%). NCDS refers to the 1958 National Child Development Study; BCS70 refers to the 1970 British Cohort study; NS refers to the 1989-90 Next Step study; MCS refers to the 2000-01 Millennium Cohort Study.

Very little variation was found across the different aged cohorts. Just over a quarter of the sample reported being financially worse off compared to before the pandemic (27% - which varied between 26%-29% across all four cohorts) and 14% of the sample made new benefit claims during the pandemic (which varied between 13%-16% across the cohorts). Of those who reported income both before and after the pandemic, 18% reported a decrease of greater than or equal to 5% (this varied between 17%-19% across the cohorts).

### Financial wellbeing

Long COVID was associated with reporting being financially worse off compared to pre-pandemic (Figure 1). The risk of being financially worse off increased with the length of time that individuals were unable to function as normal due to COVID-19. Relative to those that had not contracted SARS-CoV-2, the risk of being financially worse off was greatest among those with COVID-19 symptoms that lasted longer than 12 weeks (adjusted RRR=1.85, 95% CI=1.43, 2.41).

**Figure 1:**
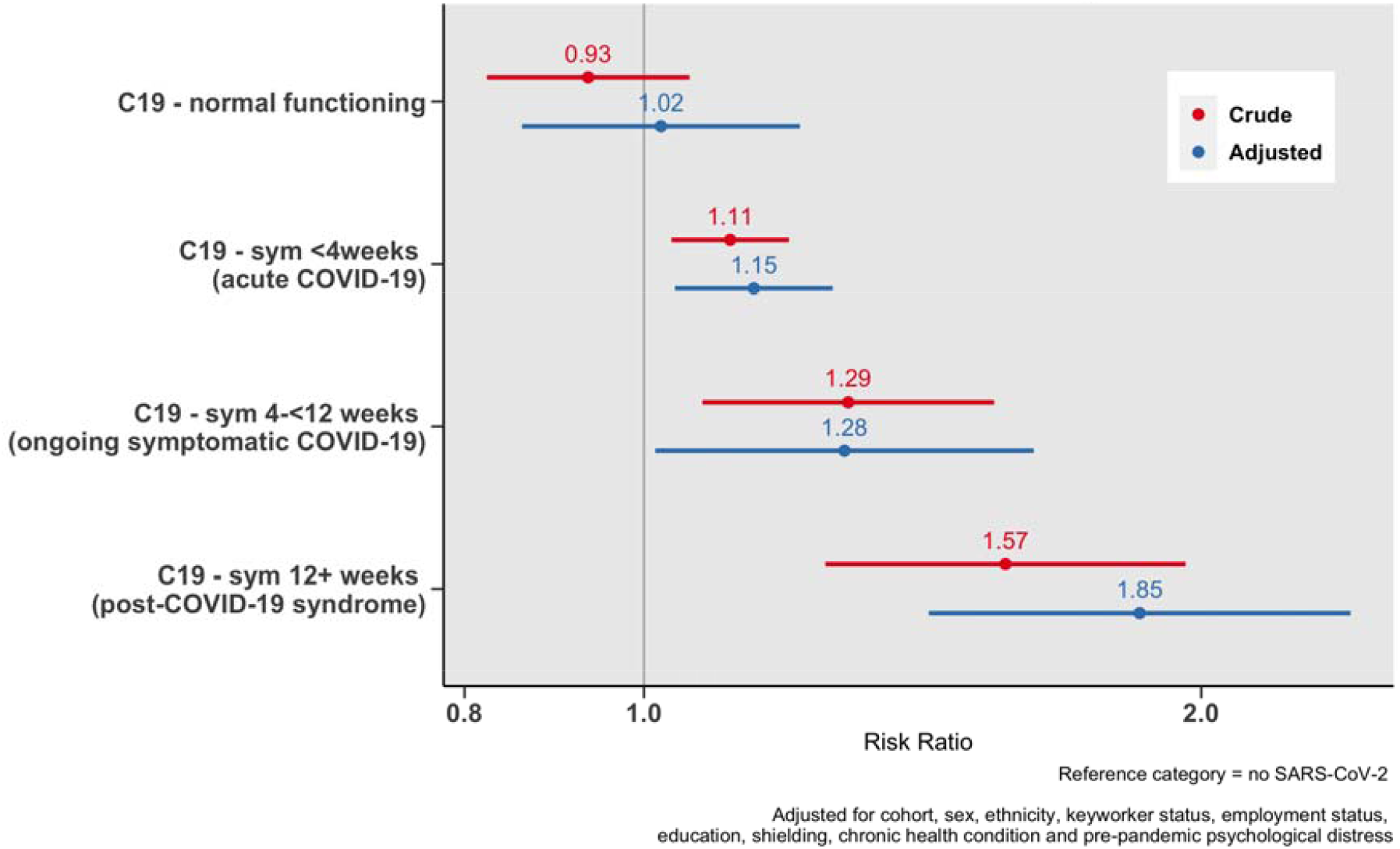
Association between duration of COVID-19 symptoms and financial wellbeing.

### New benefit claims

Long COVID was associated with new benefit claims made during the pandemic, with the greatest risk among those with COVID symptoms lasting >12 weeks (adjusted RRR=1.81, 95% CI=1.22, 2.70 – see Figure 2), relative to those who had not contracted SARS-CoV-2. The most common benefit claims for those with long COVID were Universal Credit (37%) followed by the self-employed COVID support scheme (27% - see Supplementary Material for more information on new benefit claims).

**Figure 2:**
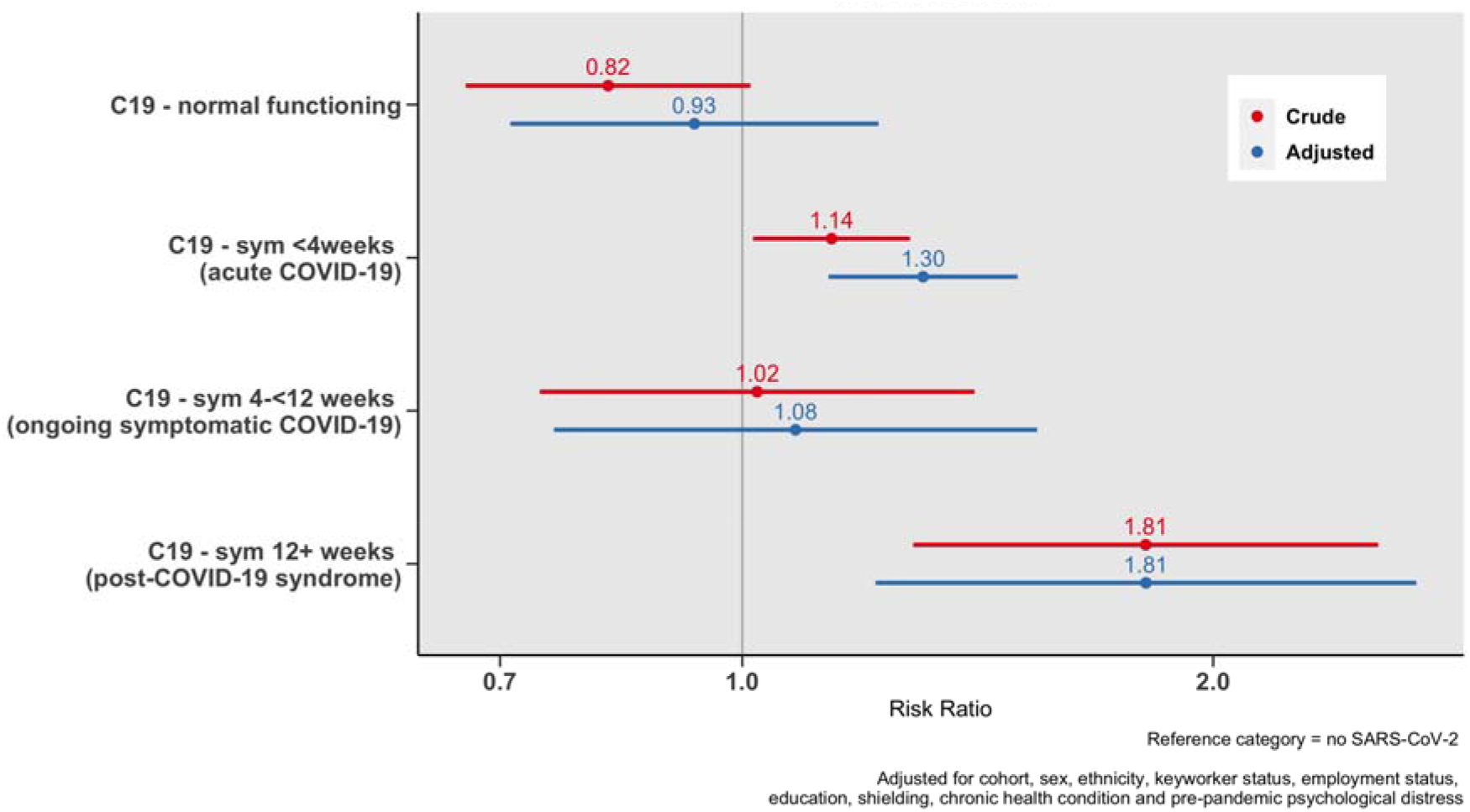
Association between COVID severity and new benefit claims.

### Change in household income during the pandemic

Figure 3 shows the associations between a four-category measure of long COVID and a decrease in weekly household income by at least 5%. Findings suggest that those with COVID-19 symptoms which last longer than 4 weeks have a greater risk of experiencing a decrease in income in the adjusted model. However, due to lack of power and resulting wide confidence intervals, our data are also compatible with the null hypothesis of no association (adjusted RRR=1.23, CI=0.93, 1.63).

**Figure 3:**
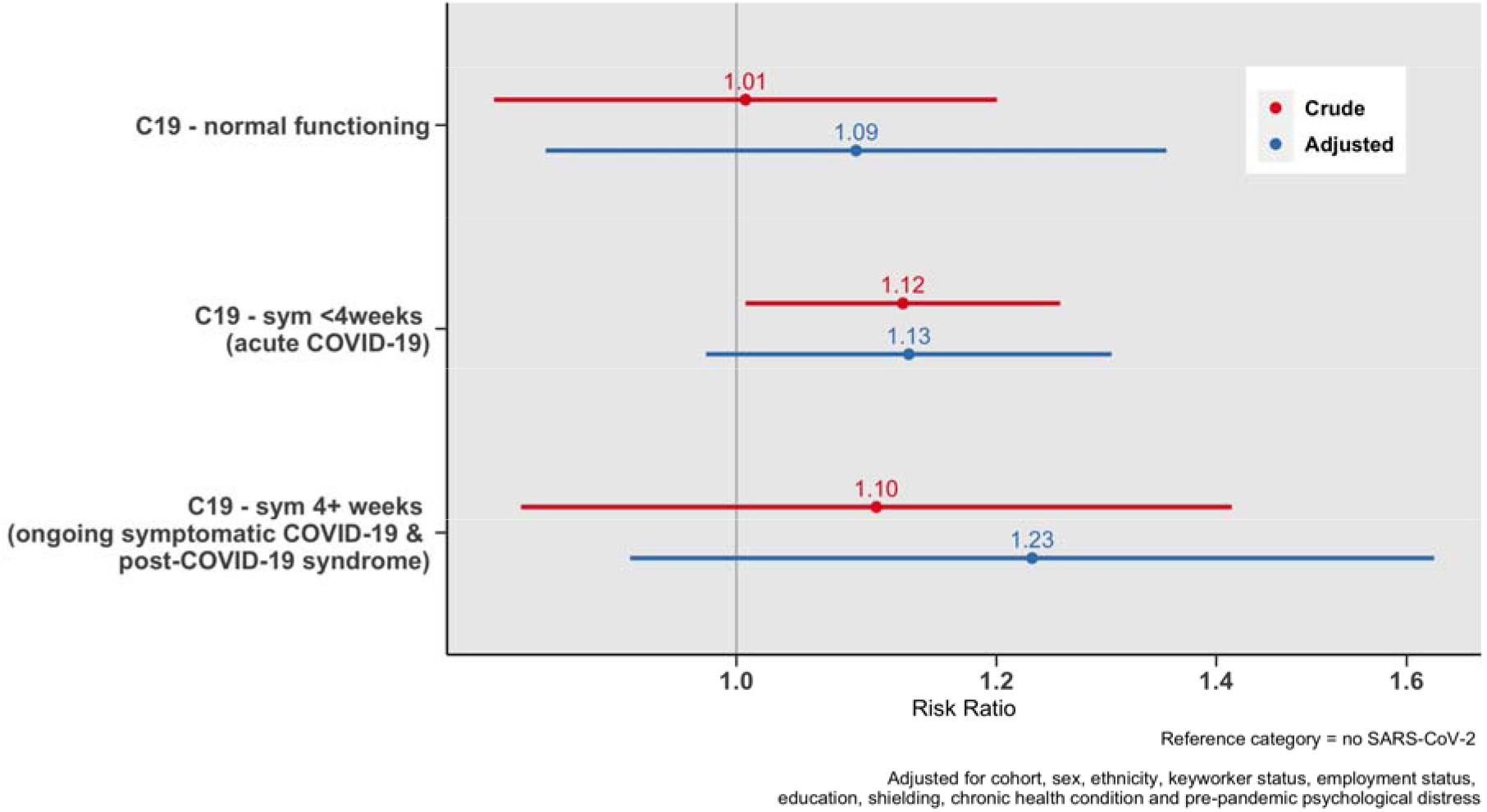
Association between duration of COVID-19 symptoms and a decrease in weekly household income of at least 5%.

Adjusted analysis conducted on imputed data produced findings that did not meaningfully differ from weighted analysis (see Supplementary Material). This was also the case for financial wellbeing and new benefit claims outcomes.

### Stratified analysis

The association between long COVID and financial disruption was modified by sex but not education.. The primary difference between male and female participants was that, for those with symptoms between 4 and 12 weeks, male participants were more likely to report being financially worse off while female participants were not. For male participants, those with symptoms lasting longer than 4 weeks report being financially worse off compared to those who had not contracted COVID-19. While for female participants, only those with symptoms lasting longer than 12 weeks reported being financially worse off compared to those who had not contracted COVID-19. No further evidence of effect modification was detected for other outcomes (see Supplementary Material for results of stratified analysis).

### Sensitivity analysis

Analyses were re-weighted using population age composition weights to represent each cohort based on the actual distribution in the overall population. Highly similar estimates to the main analysis were found. Analysis of subjective financial wellbeing and benefit claim outcomes was also conducted using a four-category measure of long COVID where symptoms lasting between 4-12 weeks (ongoing symptomatic COVID-19) and 12+ weeks (post-COVID-19 syndrome) were combined into a single category (symptoms 4+ weeks). This sensitivity analysis was conducted to increase statistical power (as relatively low cell counts were seen in the post-COVID-19 syndrome category), demonstrating consistent findings to the main analyses. Finally, the exclusion of 996 participants who reported infection <12 weeks prior to the 3^rd^ wave did not alter findings reported in the main results section.

Additional analysis was conducted to examine associations between OECD equivalised weekly household income during the pandemic (adjusting for retrospective pre-pandemic weekly household income) and a four-category measure of COVID severity. Results from this analysis suggest that those with COVID-19 symptoms which last longer than 4 weeks have decreased household income (see Supplementary Material for further details of sensitivity analyses).

## Discussion

Using data from four representative UK birth cohorts, we found that long COVID is associated with financial disruption after adjusting for a wide range of potential confounders, including pre-pandemic factors. COVID-19 with symptoms that impeded the ability to function normally was reported by 13% of the participants with 0.6% reporting such symptoms for at least 12 weeks. Long COVID is associated with deteriorating subjective financial well-being (particularly for those with post-COVID syndrome). Post-COVID syndrome is also associated with new benefit claims and potentially a decrease in household income. Associations were broadly similar in different sexes and highest education groups. These results were not meaningfully altered when scaled to represent the population by age, nor after several sensitivity analyses.

These findings contribute to a growing body of urgently needed research. Our estimation of the UK prevalence of people with either ongoing symptomatic COVID-19 or post-COVID-19 syndrome is 12.5% which is in line with some UK estimates (8). Though the prevalence of post-COVID-19 syndrome specifically was very low (0.6% of our sample), in the UK, this translates to over 400,000 people. This is concerning given that we found that those with post-COVID-19 syndrome had the worst financial outcomes compared to those who had not contracted COVID-19.

Our findings are also in agreement with similar UK studies which have found evidence to suggest that long COVID may be associated with financial disruption (21, 24). Any negative financial outcomes experienced by those with post-COVID-19 syndrome will almost certainly impact other household members who may be required to take on additional caring responsibilities and therefore require reduced / flexible working hours as well as financial compensation in the form of benefits or other forms of government support. Healthcare and financial support for those with long COVID and their families will likely be long-term also considering the possibility of acquiring a new disability (22) (which can also carry a financial penalty and place further burden on healthcare systems).

### Strengths and limitations

This study conducted an analysis of the financial impact of long COVID using data from four population-based UK birth cohorts capitalising on their rich pre-pandemic longitudinal data that allowed us to adjust for a wide range of potential confounders. Despite this, and as in any observational study, bias due to residual confounding is possible. Although individuals with minimal or no symptoms were considered as the control group in our analysis (42), the absence of any observed association between this group and our outcomes may suggest the presence of limited unmeasured confounding variables. While this provides some degree of reassurance, it is important to acknowledge that potential biases cannot be fully eliminated. Our data come from surveys during the COVID-19 pandemic where non-response bias combined with selective attrition may have biased results. We employed multiple imputation and inverse probability weighting (34) to restore sample representativeness. Long COVID and our outcomes were self-reported, measurement error due to self-report bias is another limitation that could have influenced our findings.

Long COVID in our data was only captured during the first year of the pandemic (until Feb/March 2021). As a result, relatively few people reported long COVID which meant some analyses may have been underpowered despite our efforts to combine multiple cohorts. Though to address this, multiple sensitivity analyses (that returned similar results to our main analysis) were carried out. Currently, cohort studies are collecting data on long COVID so that future studies can achieve a fuller assessment of its ongoing financial impacts (22). This study also adopts retrospective estimates of pre-COVID-19 household income which may have been subject to bias. Though pre-pandemic measures of household income were available to use in previous waves of each respective cohort, these waves took place between 2013-2018 (NCDS = 2013, NS = 2015, BCS = 2016, MCS = 2018) and therefore would likely have not reflected the actual household income of participants immediately prior to the pandemic.

We find evidence that long COVID is associated with being financially worse off but only suggestive evidence to support a link between long COVID and change in household income. This may be due to lack of power, indeed, the coefficients for change in household income seem consistent with the subjective measure. Furthermore, any impact on income may have been – at least partially - offset by increased use of benefits. Finally, our measure of financial well-being is subjective and therefore subject to self-report bias but has the added benefit of revealing (to some extent at least) a population’s confidence in the prospect of economic circumstances. It also indirectly accounts for expenses, and therefore, in some ways, can give a more nuanced understanding of financial coping than household income. Indeed, subjective, as opposed to objective, financial well-being, may be more relevant in certain contexts, particularly as a driver of poor mental health (43).

### Implications and future research

This research presents an initial inspection of the short-term financial impact of long COVID at an individual level during the first year of the pandemic. Considering the well-known link between financial circumstances and health (44, 45), financial disruption due to long COVID may exacerbate existing health inequalities, which will be in addition to the direct health impact of long COVID (46). Further research is needed to expand our understanding of the potential impact of long COVID on employment and financial outcomes for individuals directly affected but also on employers and the economy (24). Data currently being gathered by ongoing UK cohorts and other longitudinal population surveys in conjunction with linkages with electronic health records and routinely collected administrative data should be employed to further understand the medium-term social, economic and health impacts of long COVID.

With an estimated 200 million individuals affected globally, the potential impact of long COVID on population health and the labour force could be substantial (3). Given the scale of the issue and the potential financial implications highlighted in this research, it is imperative that those affected are provided proper health, social, and economic protections. Therefore, government-level intervention may be required. (47).

## Conclusion

This study has found evidence to suggest that long COVID can lead to worsening individual finances in the UK. If our findings reflect causal effects, more should be done to extend employment protection and financial support offered to those suffering from long COVID. Financial support from the government is made more urgent for those suffering from long COVID given the current cost of living crisis in the UK (48).

## Data Availability

Data for NCDS (SN 6137), BCS70 (SN 8547), MCS (SN 2000031) Next Steps (SN 5545),and all four COVID-19 surveys (SN 8658) are available through the UK Data Service.

## Acknowledgements

This work was supported by the National Core Studies, an initiative funded by UKRI, NIHR and the Health and Safety Executive. The COVID-19 Longitudinal Health and Wellbeing National Core Study was funded by the Medical Research Council (MC_PC_20030). MCS, NS, NCDS and BCS are supported by the Centre for Longitudinal Studies, Resource Centre 2015-20 grant (ES/M001660/1) and a host of other co-founders. The COVID-19 data collections in these four cohorts were funded by the UKRI grant Understanding the economic, social and health impacts of COVID-19 using lifetime data: evidence from 5 nationally representative UK cohorts (ES/V012789/1). RJSh, ED and SVK acknowledge funding from the Medical Research Council (MC_UU_00022/2) and the Scottish Government Chief Scientist Office (SPHSU17). SVK additionally acknowledges funding from the European Research Council (949582). Jacques Wels is funded by the Belgian National Scientific Fund (FNRS) Research Associate Fellowship (CQ) n° 40010931.

