## Supplementary Material for "Long COVID and financial outcomes: Evidence from four longitudinal population surveys"

### OECD equivalised measure of household income

Both pandemic and pre-pandemic income measures were equivalised using the Organisation for Economic Co-operation and Development (OECD) equivalence scale where household income was divided by the square root of the household size. This implies that, for example, the needs of a household of four are twice as large as one composed of a single person. Equivalised income measures were then log-transformed to account for the skewed distribution of data. Linear regression was used to examine the association between long COVID and change in weekly household income.

Additional analysis of the associations between OECD equivalized weekly household income (adjusting for retrospective pre-pandemic income) and a four-category measure of COVID severity are shown in Figure a. Findings support those reported in Figure 3, suggesting that those with COVID-19 symptoms which last longer than 4 weeks have decreased household income (adjusted β=-0.05, CI=-0.10, -0.01).

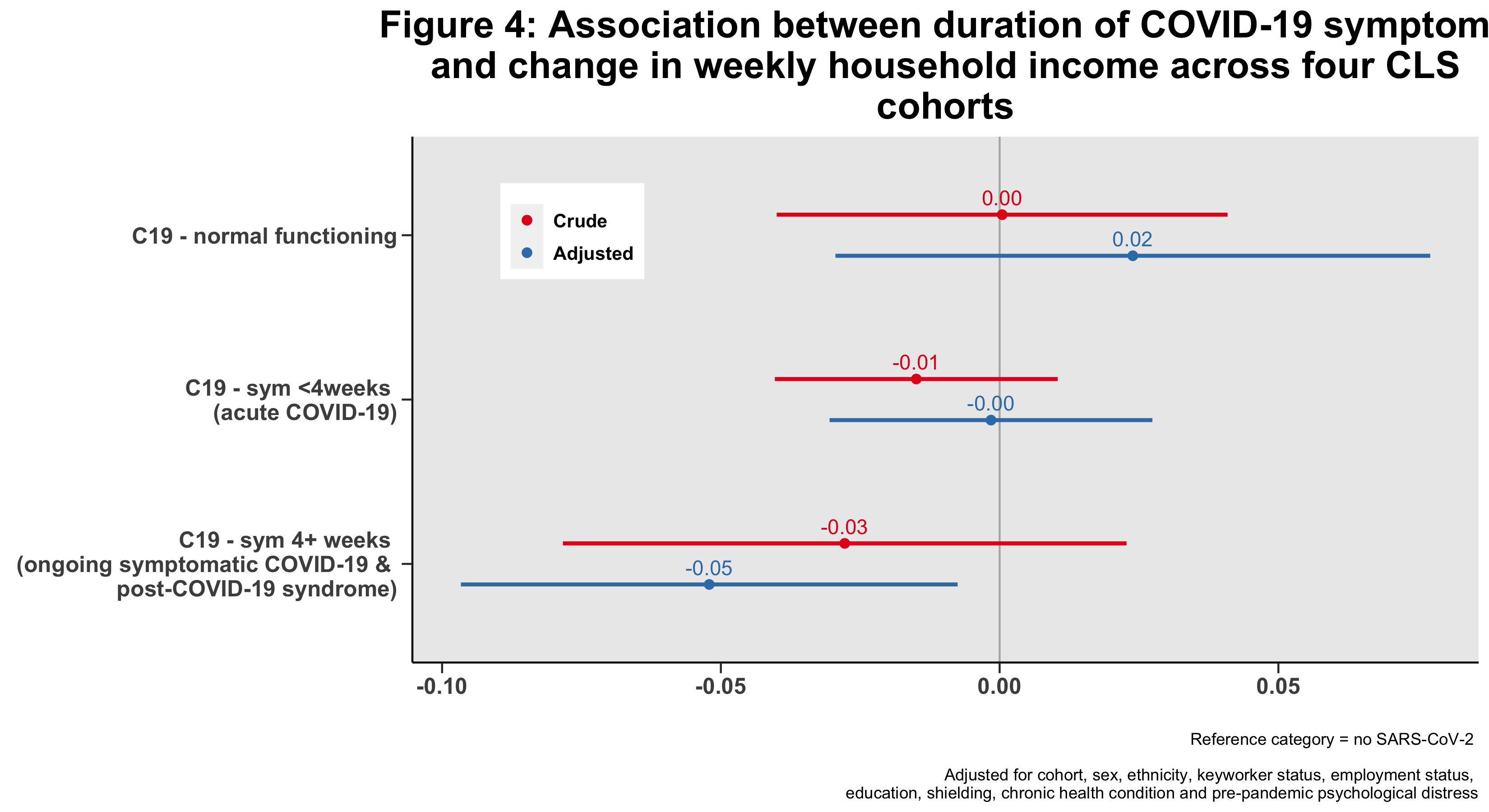

Figure a: Association between duration of COVID-19 symptoms and change in weekly household income across four CLS cohorts

### Potential confounders and DAG

**Potential confounders**

| **Characteristic** | **Overall = 20112^1^** | **NCDS N = 6467^1^** | **BCS70 N = 5421^1^** | **NS N = 4005^1^** | **MCS N = 4219^1^** |
| --- | --- | --- | --- | --- | --- |
| **Sex** |  |  |  |  |  |
| Male | 8,460 (42.2%) | 2,995 (46.3%) | 2,293 (42.3%) | 1,501 (38.0%) | 1,671 (40.1%) |
| Female | 11,652 (57.8%) | 3,472 (53.7%) | 3,128 (57.7%) | 2,504 (62.0%) | 2,548 (59.9%) |
| **Ethnicity** |  |  |  |  |  |
| White | 18,162 (94.7%) | 6,467 (100.0%) | 5,421 (100.0%) | 2,941 (90.3%) | 3,333 (85.2%) |
| Non-White | 1,950 (5.3%) | 0 (0.0%) | 0 (0.0%) | 1,064 (9.7%) | 886 (14.8%) |
| **Pre-pandemic employment** |  |  |  |  |  |
| Employed | 10,781 (61.8%) | 3,238 (54.9%) | 3,967 (86.7%) | 2,718 (84.9%) | 858 (25.0%) |
| Unemployed | 541 (2.9%) | 125 (2.1%) | 93 (2.0%) | 133 (3.2%) | 190 (4.6%) |
| Economically Inactive | 6,157 (35.3%) | 2,535 (43.0%) | 518 (11.3%) | 388 (11.8%) | 2,716 (70.4%) |
| Unknown | 2,680 | 569 | 843 | 792 | 476 |
| **Shielding during the pandemic** |  |  |  |  |  |
| No | 19,023 (94.7%) | 5,958 (92.2%) | 5,108 (94.2%) | 3,855 (96.4%) | 4,102 (97.2%) |
| Yes | 1,081 (5.3%) | 506 (7.8%) | 312 (5.8%) | 150 (3.6%) | 113 (2.8%) |
| Unknown | 8 | 3 | 1 | 0 | 4 |
| **Keyworker during the pandemic** |  |  |  |  |  |
| No | 12,521 (65.6%) | 4,422 (72.1%) | 2,709 (52.9%) | 2,019 (52.5%) | 3,371 (83.2%) |
| Yes | 6,515 (34.4%) | 1,708 (27.9%) | 2,409 (47.1%) | 1,748 (47.5%) | 650 (16.8%) |
| Unknown | 1,127 | 337 | 303 | 254 | 233 |
| **Pre-pandemic education (NVQ)** |  |  |  |  |  |
| None | 990 (5.5%) | 383 (6.0%) | 363 (7.1%) | 113 (3.2%) | 131 (4.6%) |
| NVQ1 level | 1,247 (7.3%) | 602 (9.4%) | 349 (6.8%) | 199 (5.6%) | 97 (4.9%) |
| NVQ2 level | 4,067 (24.2%) | 1,567 (24.5%) | 1,322 (25.9%) | 714 (20.6%) | 464 (25.3%) |
| NVQ3 level | 3,026 (17.8%) | 1,154 (18.1%) | 753 (14.8%) | 824 (22.7%) | 295 (15.5%) |
| NVQ4 level | 6,084 (36.3%) | 2,349 (36.7%) | 1,878 (36.9%) | 1,100 (30.9%) | 757 (43.5%) |
| NVQ5 level | 1,540 (9.0%) | 337 (5.3%) | 431 (8.5%) | 664 (17.1%) | 108 (6.2%) |
| Unknown | 3,351 | 75 | 325 | 405 | 2,545 |
| **Chronic health condition (pre-pandemic)** |  |  |  |  |  |
| No | 13,586 (72.9%) | 4,122 (68.1%) | 3,161 (64.6%) | 2,932 (80.0%) | 3,371 (82.2%) |
| Yes | 5,072 (27.1%) | 1,934 (31.9%) | 1,730 (35.4%) | 686 (20.0%) | 722 (17.8%) |
| Unknown | 1,477 | 411 | 530 | 402 | 134 |
| **Probable psychological distress (pre-pandemic)** |  |  |  |  |  |
| No | 15,268 (82.7%) | 5,218 (87.2%) | 4,086 (83.5%) | 2,627 (74.6%) | 3,337 (82.8%) |
| Yes | 3,212 (17.3%) | 769 (12.8%) | 805 (16.5%) | 919 (25.4%) | 719 (17.2%) |
| Unknown | 1,654 | 480 | 530 | 464 | 180 |
| ^1^n_unweighted (%) | | | | | |

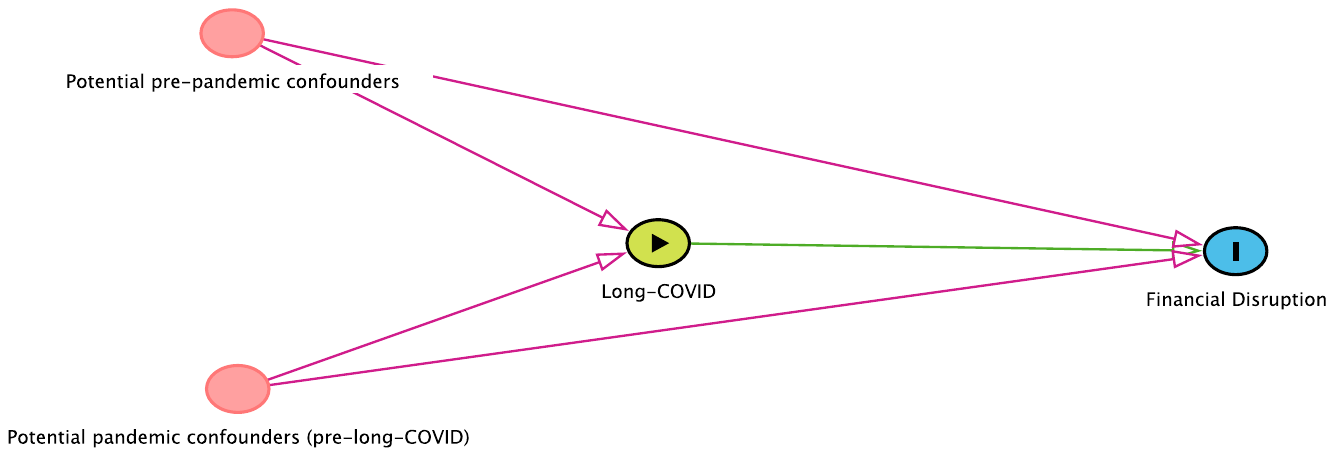

**Figure b: Directed acyclic graph (DAG) to illustrate confounders relationship with exposure and outcomes**

### Re-weighted analysis using scaled populations weights

**Long-COVID associated with subject of financial well-being, new benefit claims and change in weekly household income (population age composite weights)**

|  | **Crude estimates using population age composition weights** |
| --- | --- |
| **Financial coping** | **RRR (95% CI)** |
| C19 - normal functioning | 0.93 (0.82, 1.06) |
| C19 - symptoms <4weeks | 1.11 (1.03, 1.20) |
| C19 - symptoms 4-<12 weeks | 1.29 (1.06, 1.56) |
| C19 - symptoms 12+ weeks | 1.52 (1.20, 1.93) |
| **New benefit claims** |  |
| C19 - normal functioning | 0.83 (0.67, 1.03) |
| C19 - symptoms <4weeks | 1.19 (1.06, 1.34) |
| C19 - symptoms 4-<12 weeks | 1.16 (0.85, 1.59) |
| C19 - symptoms 12+ weeks | 1.84 (1.29, 2.62) |
| **Decrease in weekly household income** ≥ **5%** | |
| C19 - normal functioning | 0.92 (0.79, 1.06) |
| C19 - symptoms <4weeks | 1.00 (0.91, 1.10) |
| C19 - symptoms 4+ weeks | 1.04 (0.84, 1.29) |

### Complete case analysis and imputed data

**Table C1: Long-COVID associated with subject of financial well-being and new benefit claims (complete case and MI)**

| **Long-COVID** | **Crude** | | **Adjusted** | | **Adjusted (MICE Imputed data)** |
| --- | --- | --- | --- | --- | --- |
|  | **n** | **RRR (95% CI)** | **n** | **RRR (95% CI)** | **RRR (95% CI)** |
| **Financial wellbeing** |  |  |  |  |  |
| no covid | 16,731 | -- | 11,323 | -- | -- |
| C19 - normal functioning | 879 | 0.93 (0.82, 1.06) | 505 | 1.02 (0.86, 1.21) | 0.96 (0.85, 1.09) |
| C19 - symptoms <4weeks | 2,139 | 1.11 (1.03, 1.20) | 1,326 | 1.15 (1.04, 1.26) | 1.13 (1.05, 1.22) |
| C19 - symptoms 4-<12 weeks | 251 | 1.29 (1.08, 1.55) | 176 | 1.28 (1.01, 1.62) | 1.28 (1.07, 1.54) |
| C19 - symptoms 12+ weeks | 112 | 1.57 (1.25, 1.96) | 78 | 1.85 (1.43, 2.41) | 1.62 (1.30, 2.03) |
| **New benefit claims** |  |  |  |  |  |
| no covid | 16,731 | -- | 14,314 | -- | -- |
| C19 - normal functioning | 879 | 0.85 (0.69, 1.04) | 638 | 0.94 (0.72, 1.22) | 0.92 (0.75, 1.13) |
| C19 - symptoms <4weeks | 2,139 | 1.19 (1.06, 1.33) | 1,670 | 1.34 (1.17, 1.53) | 1.21 (1.08, 1.35) |
| C19 - symptoms 4-<12 weeks | 251 | 1.12 (0.83, 1.51) | 233 | 1.16 (0.83, 1.63) | 1.04 (0.78, 1.40) |
| C19 - symptoms 12+ weeks | 112 | 1.79 (1.27, 2.53) | 99 | 1.79 (1.20, 2.66) | 1.71 (1.22, 2.39) |

**Long-COVID associated with a decrease in weekly**

**household income** ≥ **5% (complete case and MI)**

| **Long-COVID** | **Crude** | | **Adjusted** | |
| --- | --- | --- | --- | --- |
|  | **RRR (95% CI)** | | **RRR (95% CI)** | |
| **Decrease in weekly household income** ≥ **5% (complete case)** | | | | |
| C19 - normal functioning | 0.91 (0.79, 1.05) | | 1.03 (0.85, 1.23) | |
| C19 - symptoms <4weeks | 1.00 (0.91, 1.09) | | 1.05 (0.93, 1.18) | |
| C19 - symptoms 4+ weeks | 1.07 (0.88, 1.30) | | 1.25 (1.02, 1.53) | |
| **Decrease in weekly household income** ≥ **5% (missing data imputed)** | | | | |
| C19 - normal functioning | | 1.01 (0.95, 1.08) | | 1.02 (0.95, 1.09) |
| C19 - symptoms <4weeks | | 1.01 (0.96, 1.07) | | 1.02 (0.97, 1.07) |
| C19 - symptoms 4+ weeks | | 1.05 (0.97, 1.14) | | 1.07 (0.99, 1.15) |

### Sensitivity analysis using a four-category measure of long-COVID & stratified analysis

**Table E1: Long-COVID (four-category) associated with financial wellbeing**

| **Financial wellbeing** | | |
| --- | --- | --- |
| **Long-COVID** | **Crude** | **Adjusted** |
|  | **RRR (95% CI)** | **RRR (95% CI)** |
| C19 - normal functioning | 0.93 (0.82, 1.06) | 0.95 (0.81, 1.11) |
| C19 - symptoms <4weeks | 1.11 (1.03, 1.20) | 1.11 (1.02, 1.21) |
| C19 - symptoms 4+weeks | 1.38 (1.19, 1.59) | 1.32 (1.12, 1.56) |

**Table E2: Long-COVID (four-category) associated with new benefit claims**

| **New benefit claims** | | |
| --- | --- | --- |
| **Long-COVID** | **Crude** | **Adjusted** |
|  | **RRR (95% CI)** | **RRR (95% CI)** |
| C19 - normal functioning | 0.85 (0.69, 1.04) | 0.94 (0.72, 1.22) |
| C19 - symptoms <4weeks | 1.19 (1.06, 1.33) | 1.34 (1.17, 1.53) |
| C19 - symptoms 4+weeks | 1.33 (1.06, 1.68) | 1.36 (1.05, 1.77) |

**Long-COVID associated with subjective financial well-being, stratified by sex**

| **Financial wellbeing** |  | | |  | | |
| --- | --- | --- | --- | --- | --- | --- |
| **Long-COVID** | **Male** | | | **Female** | | |
|  | **n** | **RRR** | **95% CI** | **n** | **RRR** | **95% CI** |
| no covid | 7,362 | — | — | 9,971 | — | — |
| C19 - normal functioning | 462 | 0.83 | 0.68, 1.02 | 469 | 1.04 | 0.88, 1.21 |
| C19 - symptoms <4weeks | 851 | 1.10 | 0.97, 1.24 | 1,372 | 1.12 | 1.02, 1.23 |
| C19 - symptoms 4-<12 weeks | 77 | 1.91 | 1.50, 2.42 | 178 | 1.02 | 0.80, 1.32 |
| C19 - symptoms 12+ weeks | 43 | 1.58 | 1.07, 2.32 | 74 | 1.56 | 1.18, 2.05 |

### New benefit claims

|  | **Overall = 20112^1^** | **no covid N = 16731^1^** | **C19 - normal functioning N = 879^1^** | **C19 - sym <4weeks N = 2139^1^** | **C19 - 4+ weeks N = 363^1^** |
| --- | --- | --- | --- | --- | --- |
| **New benefit claims?** |  |  |  |  |  |
| No | 17,256 | 14,410 (83.33%) | 770 (4.59%) | 1,780 (10.37%) | 296 (1.70%) |
| Yes | 2,856 | 2,321 (81.52%) | 109 (3.71%) | 359 (12.43%) | 67 (2.34%) |
| ^1^n_unweighted (%) | | | | | |

|  | **Free school dinners** | **Universal credit** | **Employment support** | **Sick pay** | **Council tax support** | **COVID-19 self-employment income support** | **Career allowance** | **Test and trace** |
| --- | --- | --- | --- | --- | --- | --- | --- | --- |
| no covid | 79 (3.4%) | 873 (37.6%) | 249 (10.7%) | 199 (8.6%) | 281 (12.1%) | 771 (33.2%) | 302 (13.0%) | 32 (1.4%) |
| C19 - normal functioning | 4 (3.7%) | 41 (37.6%) | 7 (6.4%) | 17 (15.6%) | 11 (10.1%) | 31 (28.4%) | 4 (3.7%) | 9 (8.3%) |
| C19 - symptoms | 14 (3.3%) | 157 (36.9%) | 39 (9.2%) | 72 (16.9%) | 56 (13.1%) | 108 (25.4%) | 40 (9.4%) | 32 (7.5%) |

|  | **Free school dinners** | **Universal credit** | **Employment support** | **Sick pay** | **Council tax support** | **COVID-19 self-employment income support** | **Career allowance** | **Test and trace** |
| --- | --- | --- | --- | --- | --- | --- | --- | --- |
| no covid | 79 (3.4%) | 873 (37.6%) | 249 (10.7%) | 199 (8.6%) | 281 (12.1%) | 771 (33.2%) | 302 (13.0%) | 32 (1.4%) |
| C19 - normal functioning | 4 (3.7%) | 41 (37.6%) | 7 (6.4%) | 17 (15.6%) | 11 (10.1%) | 31 (28.4%) | 4 (3.7%) | 9 (8.3%) |
| C19 - sym <4weeks | 13 (3.6%) | 135 (37.6%) | 32 (8.9%) | 54 (15.0%) | 46 (12.8%) | 99 (27.6%) | 27 (7.5%) | 26 (7.2%) |
| C19 - sym 4-<12 weeks | 1 (2.6%) | 12 (30.8%) | 4 (10.3%) | 10 (25.6%) | 5 (12.8%) | 4 (10.3%) | 8 (20.5%) | 5 (12.8%) |
| C19 - sym 12+ weeks |  | 10 (35.7%) | 3 (10.7%) | 8 (28.6%) | 5 (17.9%) | 5 (17.9%) | 5 (17.9%) | 1 (3.6%) |
